# Nomogram-Based Prognostic Model to predict the High blood pressure in Children and Adolescents —— Finding from 342,736 individual in China

**DOI:** 10.1101/2021.08.24.21262545

**Authors:** Jing-hong Liang, Yu Zhao, Yi-can Chen, Shan Huang, Shu-xin Zhang, Nan Jiang, Aerziguli Kakaer, Ya-jun Chen

**Author notes:** Corresponding Author: Ya-jun Chen, MD, School of public Health, Medical College of Sun Yat-Sen University, No.74 Zhongshan 2nd Road, Yuexiu District, Guangzhou, China 510080. **E-mail:** Jing-hong Liang,; Yu Zhao,; Yi-can Chen,; Shan Huang,; Nan Jiang,; Shu-xin Zhang,; Aerziguli Kakaer,.

## Abstract

**Background:** Predicting the potential risk factors of High blood pressure(HBP) among children and adolescents is still a knowledge gap. Our study aimed to establish and validate a nomogram-based model for identifying children and adolescents at risk of developing HBP based on a population-based prospective study.

**Methods:** Hypertension was defined as systolic blood pressure or diastolic blood pressure above 95^th^ percentile, using age, gender and height-specific cut-points. Penalized regression with Lasso was used to identify the strongest predictors of hypertension. Internal validation was conducted by 5-fold cross-validation and bootstrapping approach. The predictive variables were identified along with the advanced nomogram plot by conducting univariate and multivariate logistic regression analyses. A nomogram was constructed by training group comprised of 239,546(69.89%)participants and subsequently validated by externally group with 103,190(30.11%)participants.

**Results:** Of 342,736 children and adolescents, a total of 55,480(16.19%) youths were identified with HBP with mean age 11.51±1.45 year and 183,487 were boys(53.5%). Nine significant relevant predictors were identified including: age, gender, weight status, birthweight, breastfeeding, gestational hypertension, family history of obesity, family history of hypertension and physical activity. An acceptable discrimination[Area under the receiver operating characteristic curve(AUC):0.742(Development group), 0.740(Validation group)] and good calibration(Hosmer and Lemeshow statistics, P > 0.05) were observed in our models. An available web-based nomogram was built online.

**Conclusions:** This model composed of age, gender, early life factors, family history of disease, and lifestyle factors may predict the risk of HBP among children and adolescents, which has developed a promising nomogram that may aid in more accurately for identifying the HBP among youths in primary care.

**Funding Sources:** The work was supported by the National Natural Science Foundation of China (No. 81673193).

## Introduction

High blood pressure(HBP) whether in primary or secondary, were occur with 2-3% incidence in the general children(Chiolero, Cachat, Burnier, Paccaud, & Bovet, 2007; McNiece et al., 2007) and deemed as the most common and potentially reversible early marker for the development of metabolic diseases, neurological disorders, persistent of hypertension in adulthood, cardiovascular disease in particular(Lauer, Anderson, Beaglehole, & Burns, 1984; Rosner, Cook, Daniels, & Falkner, 2013), which contributing not only to global burden but also a major cause of poorer disability-adjusted life-years both for youth and adults(Collaborators, 2017; Lim et al., 2012). Of note, compared with female, male tend to have a higher proportion of arterial hypertension among 18 years old adolescents with the ratio of male to female (3-4:1)(Rosner et al., 2013). However, there are 74% children with various documented were high Blood Pressure(BP) have been underrecognized as hypertensive(Hansen et al., 2007), which likely owing to are not fully understand but are likely due to the asymptomatic presentation of hypertension, challenging patient assessments, complicated BP norms, and unclear recommendations.

Given the increasing evidence base, recommendations of identification as respect of BP in children from multiple guidelines vary somewhat(J. M. D. A et al., 2017; Dionne et al., 2016; Empar et al., 2016; None, 2017), probably attribute to differences on their region, sample size and other extrapolation of their evidence. Based on the pediatric researches, it is essential to identify inherent and exterior predictors may provide the comprehensive strategies for unraveling the pathogenesis of hypertension at individuals’ early stage.

And another concern is that the present studies yielded contradictory results on the risk factors for HBP in children. For example, some study indicate that children exposure to smoking were more likely associated with an increased risk of HBP whether in active or passive smoking(Katona et al., 2011; Kollias et al., 2009; Simonetti et al., 2011) while some reported an inconsistent evidence(Giussani, Antolini, Brambilla, Pagani, & Genovesi, 2013; Roya et al., 2016), and the meta-analysis has produced the unconvincing available evidence with the relatively low-quality epidemiologic studies(Aryanpur et al., 2019). Furthermore, previous predictive model remains inconsistent. A study performed in Switzerland identified a higher proportion of children with HBP (65%) based on the child overweight and parental HBP than another strategy in accordance with child overweight only(43%)(Bloetzer et al., 2015; Hamoen, Welten, Nieboer, Heymans, & Wang, 2020). Thus, a more plausible model to target screening for children and adolescents with HBP need to be properly addressed.

Consequently, in this study, based on the ongoing successive sectional study conducted in Guangzhou[(Guangzhou Survey on Students’ Constitution and Health), GSSCH] in 2020 with total of 1,594,048 participants, our study is aiming at assess the association of numerous prediction and hypertension and, to first ever build a nomogram statistical tool to identify the precise long-term effect modifiers of HBP in children and adolescents both by developing and internal validating the dataset established in 2020.

## Results

### Demographic characteristics

As displayed in **Supplementary Figure.1**, based on the several screening criteria, after exclusions, a total of 342,736 participants were identified. 69.89%(N=239,546)students were randomly assigned to the derivation group and 30.11% (N=103,190)to the internal validation group. Of the development group, the average age of children and adolescents was 11.51 years(2.73) with range from 6 to 18, and 11.51 years (2.74) with range from 7 to 18 in validation group. More than half of the children were boys (53.5%, n=183,487). Of 342,736 participants, 287,256(83.81%) and 55,480(16.19%) children were identified as having HBP and non-HBP, respectively.

There was existing a statistically significant difference among the majority of demographic variables in either the HBP group and non-HBP group(P<0.001), such as age, gender, birthweight and so on. We only found no significant differences in one parental education level. The demographic characteristics of the subjects was listed in **Table.1**.

**Table.1.** Comparison of baseline characteristics between HBP group and non-HBP group.

| **Characteristics** | **Total**<br>**(n= 342,736)** | **HBP**<br>**group(n=55,480)** | **Non-HBP**<br>**group(n=287,256)** | **P-value** |
| --- | --- | --- | --- | --- |
| **Age(year)(mean±SD)** | 11.51 ± 2.73 | 12.22 ± 2.88 | 11.37 ± 2.68 | < 0.001 |
| **Boys [n(%)]** | 183,487(53.5) | 35,246(63.5) | 148,241(51.6) | < 0.001 |
| **Feeding mode [n(%)]** |  |  |  |  |
| Breastfeeding | 136,442(39.8) | 21,455(38.7) | 114,987(40.0) | < 0.001 |
| Bottle feeding | 140,342(40.9) | 18,325(33.0) | 122,017(42.5) |  |
| Partial breastfeeding | 65,952(19.2) | 15,700(28.3) | 50,252(17.5) |  |
| **Birthweight (g)(mean±SD)** |  |  |  |  |
| 2500-4000 | 307,250(89.6) | 40,922(73.8) | 266,328(92.7) | < 0.001 |
| < 2500 | 34,046(9.9) | 14,117(25.4) | 19,929(6.9) |  |
| > 4000 | 1,440(0.4) | 441(0.8) | 999(0.3) |  |
| **Gestational hypertension [n(%)]** | 6,133(1.8) | 1,355(2.4) | 4,778(1.7) | < 0.001 |
| **SBP(mmHG)** | 104.05 ± 10.17 | 116.10 ± 8.25 | 101.73 ± 8.76 | < 0.001 |
| **DBP(mmHG)** | 64.95 ± 6.88 | 71.17 ± 6.48 | 63.75 ± 6.29 | < 0.001 |
| **BMI(kg/m2)(mean±SD)** | 18.00 ± 3.36 | 19.25 ± 3.88 | 17.75 ± 3.19 | < 0.001 |
| **Weight status [n(%)]** |  |  |  |  |
| Underweight | 39,382(11.5) | 8,682(15.6) | 30,700(10.7) | < 0.001 |
| Normal weight | 241,1212(704.4) | 36,145(65.1) | 205,067(71.4) |  |
| Overweight | 26,443(7.7) | 6,749(12.2) | 19,694(6.9) |  |
| Obese | 35,699(10.4) | 3,904(7.0) | 31,795(11.1) |  |
| **Parental smoking status [n(%)]** |  |  |  |  |
| Never smokers | 156,156(45.6) | 25,208(45.4) | 130,948(45.6) | < 0.001 |
| Former smokers | 14,839(12.2) | 6,987(12.6) | 34,852(12.1) |  |
| Current smokers | 144,741(42.2) | 23,285(42.0) | 121,456(42.3) |  |
| **Parental education level [n(%)]** |  |  |  | 0.552 |
| Primary school or below | 5,194(1.5) | 866(1.6) | 4,328(1.5) |  |
| Secondary school | 87,255(25.5) | 14,085(25.4) | 73,170(25.5) |  |
| Senior high school or junior school | 93,230(27.2) | 15,023(27.1) | 78,207(27.2) |  |
| Junior college or university | 142,442(41.6) | 23,084(41.6) | 119,358(41.6) |  |
| Graduate or above | 14,615(4.3) | 2,422(4.4) | 12,193(4.2) |  |
| **Family history of hypertension [n(%)]** | 126,981(37.0) | 12,593(22.7) | 114,388(39.8) | < 0.001 |
| **Family history of obesity [n(%)]** | 48,135(14.0) | 8,295(15.0) | 39,840(13.9) | < 0.001 |
| **Household monthly income [n(%)]** |  |  |  |  |
| <5000,RMB | 156,994(45.8) | 24,547(44.2) | 132,447(46.1) |  |
| 5000~7999, RMB | 78,761(23.0) | 13,161(23.7) | 65,600(22.8) | < 0.001 |
| 8000~11999, RMB | 69,654(20.3) | 11,657(21.0) | 57,997(20.2) |  |
| ≥12000, RMB | 37,327(10.9) | 6,115(11.0) | 31,212(10.9) |  |
| **PA time(Mins/day)(mean±SD)** | 62.03 ± 53.64 | 62.63 ± 54.22 | 61.92 ± 53.53 | 0.004 |
| **Average outdoor physical activity time [n(%)]** |  |  |  |  |
| <1 h/day | 157,116(45.8) | 29,274(52.8) | 127,842(44.5) |  |
| 1-1.9 h/day | 151,117(44.1) | 20,813(37.5) | 130,304(45.4) | < 0.001 |
| 2-4 h/day | 29,311(8.6) | 4,534(8.2) | 24,777(8.6) |  |
| >4 h/day | 5,192(1.5) | 859(1.5) | 4,333(1.5) |  |
| **SBT time(Mins/day)(mean±SD)** | 41.97 ± 47.85 | 44.10 ± 51.37 | 41.56 ± 47.13 | < 0.001 |
| **Average Screen-based time [n(%)]** |  |  |  |  |
| <2 h/day | 324,517(94.7) | 52,030(93.8) | 272,487(94.9) |  |
| 2-4 h/day | 14,861(4.3) | 2,761(5.0) | 12,100(4.2) | < 0.001 |
| >4 h/day | 3,358(1.0) | 689(1.2) | 2,669(0.9) |  |
| **Fried food intake (serving/week)** | 0.89 ± 1.18 | 0.91 ± 1.23 | 0.88 ± 1.17 | < 0.001 |

Data were presented as mean (SD), median (IQR) or n (%).
HBP, High blood pressure; PA, Physical activity; SBT, Screen-based time; SDP, Systolic blood pressure; DBP, Diastolic blood pressure.

The prevalence of HBP was 16.19%[Development group:16.20%(n=38,807), Validation group: 16.16%(n=16,673)] judged by the 95^th^ percentile for gender, age and height. The mean SBP and DBP were 104.10 ± 10.17 mmHg and 64.95 ± 6.88 mmHg in total participants, respectively.

### Multivariable predictors of hypertension

According to the pre-establish preoperative variables evaluated, Lasso regression was showed that there are 14 candidate predictors including age, gender, birthweight, weight status, feeding mode, gestational hypertension, parental smoking status, parental education, family history of obesity, family history of hypertension, household monthly income, average outdoor physical activity time, average screen-based time and fired food intake with a high correlated in the incidence of HBP for children and adolescents (Shown in **Figure.3**), while there are only 9 of them(age, gender, birthweight, weight status, feeding mode, gestational hypertension, family history of obesity, family history of hypertension and average outdoor physical activity time) were independently associated with a higher probability of HBP after the backward elimination procedure of multivariable logistic analysis. **Table.2** shows the results of multivariable predictors. The nomogram plot based on all the 9 independently predictors calculated by the generalized linear models was depicted in **Figure.1** with single score of each predictor which contribute to a total score for predicting HBP. The discrimination of model as per the above 9 significantly risk factors were detected by the apparent C-statistic for the model with value of 0.742 in development group and 0.740 in validation group(Shown in **Figure.2**). With the model included all the 14 candidate factors, the AUC were 0.744 and 0.741 in development and validation group, respectively(Shown in **Supplementary Figure.2)**. All of which mentioned above tests were indicated good discrimination, which shown in **Figure.2**. The Decision curve analysis(DCA) which presented in **Supplementary Figure.3** was further suggest that our nomogram yielded a high clinical net benefit, which is of great value for accurate individualized measurement of the incidence of hypertension among children and adolescents.

**Table.2.** Multivariable Logistic regression analysis of variables predicting HBP in the development Group.

| **Variables** |  | **OR** | **P-value** |
| --- | --- | --- | --- |
| **Age** |  | 1.142 (1.137 to 1.147) | <0.001 |
| **Gender** |  |  |  |
| Boy | Ref |  |  |
| Girl |  | 0.593 (0.578 to 0.607) | <0.001 |
| **Feeding mode** |  |  |  |
| Breastfeeding | Ref |  |  |
| Bottle feeding |  | 1.656 (1.608 to 1.706) | <0.001 |
| Partial breastfeeding |  | 0.833 (0.811 to 0.856) | <0.001 |
| **Birthweight** |  |  |  |
| 2500-4000 | Ref |  |  |
| < 2500 |  | 4.906 (4.757 to 5.060) | <0.001 |
| > 4000 |  | 2.494 (2.157 to 2.876) | <0.001 |
| **Gestational hypertension** |  |  |  |
| Gestational hypertension | Ref |  |  |
| Non-Gestational hypertension |  | 0.720 (0.665 to 0.779) | <0.001 |
| **Weight status** |  |  |  |
| Underweight | Ref |  |  |
| Normal weight |  | 0.715 (0.684 to 0.747) | <0.001 |
| Overweight |  | 1.825 (1.765 to 1.888) | <0.001 |
| Obese |  | 2.430 (2.337 to 2.526) | <0.001 |
| **Parental smoking status** |  |  |  |
| Never smokers | Ref |  |  |
| Former smokers |  | 1.020 (0.982 to 1.059) | 0.301 |
| Current smokers |  | 1.012 (0.987 to 1.038) | 0.362 |
| **Parental education level** |  |  |  |
| Primary school or below | Ref |  |  |
| Secondary school |  | 1.132 (1.029 to 1.247) | 0.012 |
| Senior high school or junior school |  | 1.188 (1.080 to 1.310) | <0.001 |
| Junior college or university |  | 1.352 (1.229 to 1.490) | <0.001 |
| Graduate or above |  | 1.521 (1.361 to 1.701) | <0.001 |
| **Family history of hypertension** |  |  |  |
| Non-Family history of hypertension | Ref |  |  |
| Family history of hypertension |  | 2.618 (2.545 to 2.693) | <0.001 |
| **Family history of obesity** |  |  |  |
| Family history of obesity | Ref |  |  |
| Non-Family history of obesity |  | 0.706 (0.682 to 0.731) | <0.001 |
| **Household monthly income** |  |  |  |
| <5000,RMB | Ref |  |  |
| 5000~7999, RMB |  | 1.058 (1.026 to 1.090) | <0.001 |
| 8000~11999, RMB |  | 1.045 (1.011 to 1.079) | <0.001 |
| ≥12000, RMB |  | 1.055 (1.012 to 1.100) | 0.011 |
| **Average outdoor physical activity time** |  |  |  |
| <1 h/day | Ref |  |  |
| 1-1.9 h/day |  | 0.672 (0.655 to 0.689) | <0.001 |
| 2-4 h/day |  | 0.724 (0.693 to 0.756) | <0.001 |
| >4 h/day |  | 0.755 (0.687 to 0.829) | <0.001 |
| **Average Screen-based time** |  |  |  |
| <2 h/day | Ref |  |  |
| 2-4 h/day |  | 1.053 (0.996 to 1.113) | 0.066 |
| >4 h/day |  | 1.191 (1.069 to 1.325) | <0.001 |
| **Fried food intake (serving/week)** |  | 1.035 (1.026 to 1.045) | <0.001 |

**Figure.1.**
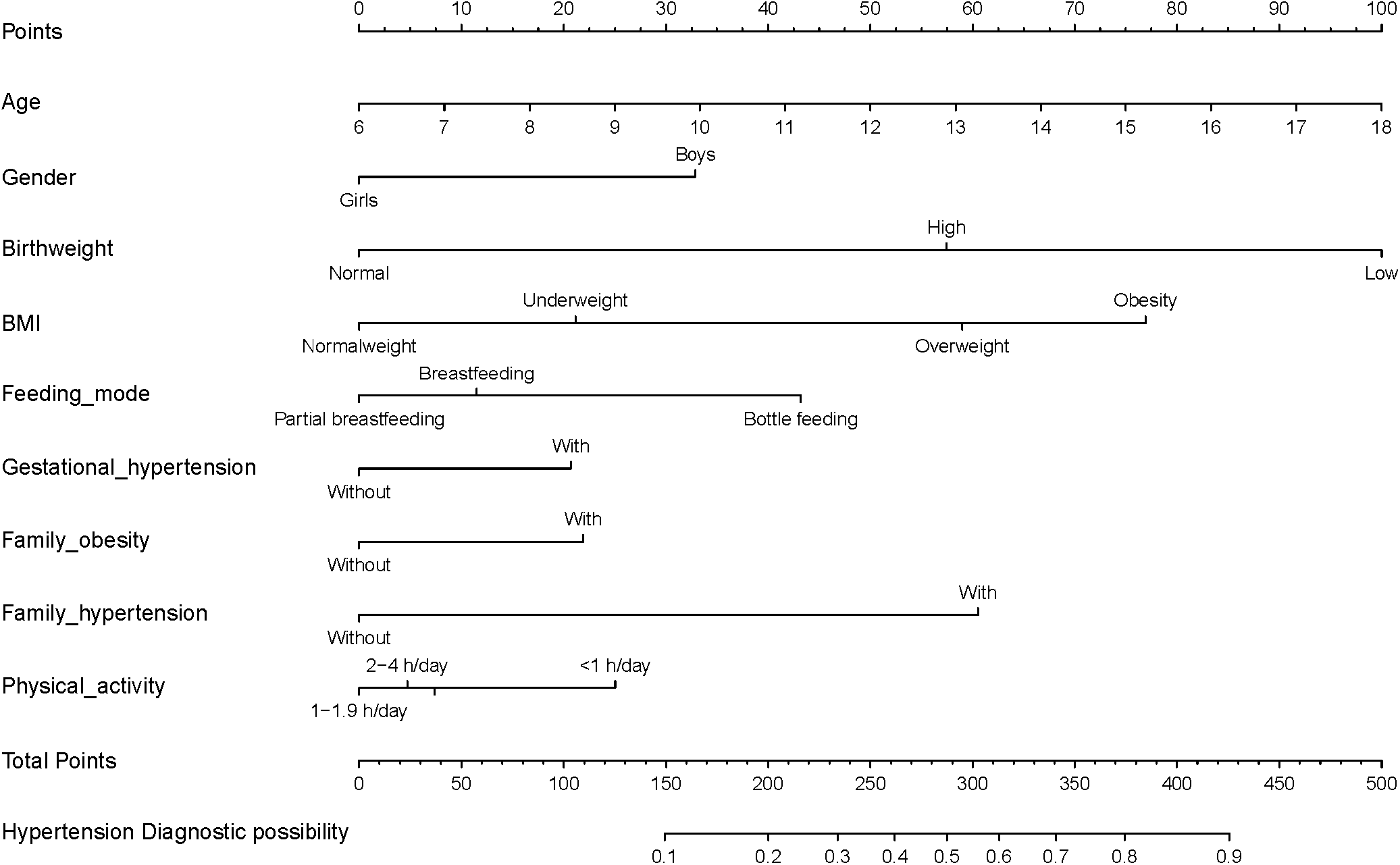
Clinical nomogram for predicting probability of developing HBP among children and adolescents, and its predictive performance. To use the nomogram, an individual HBP contact’s values is located on each variable axis, and a line is drawn downward to the risk of HBP axes to detect the hypertension probability. As an example of how this nomogram can be calculated, we can take an 18 years old obese boy who was received a bottle feeding in early life, with gestational hypertension, family of hypertension and obesity, and less than one-hour physical activity expenditure. By drawing a line up towards the points for each of the variables this student will have 100 points(Age), 32 points(Gender), 74 points(BMI status), 42 points(Feeding mode), 20 points(Gestational hypertension), 21 points(Family obesity), 59 points(Family hypertension) and 26 points(Physical activity), giving a total of 374 points(at the bottom of the figure), and a probability of HBP of 80%.

**Figure.2.**
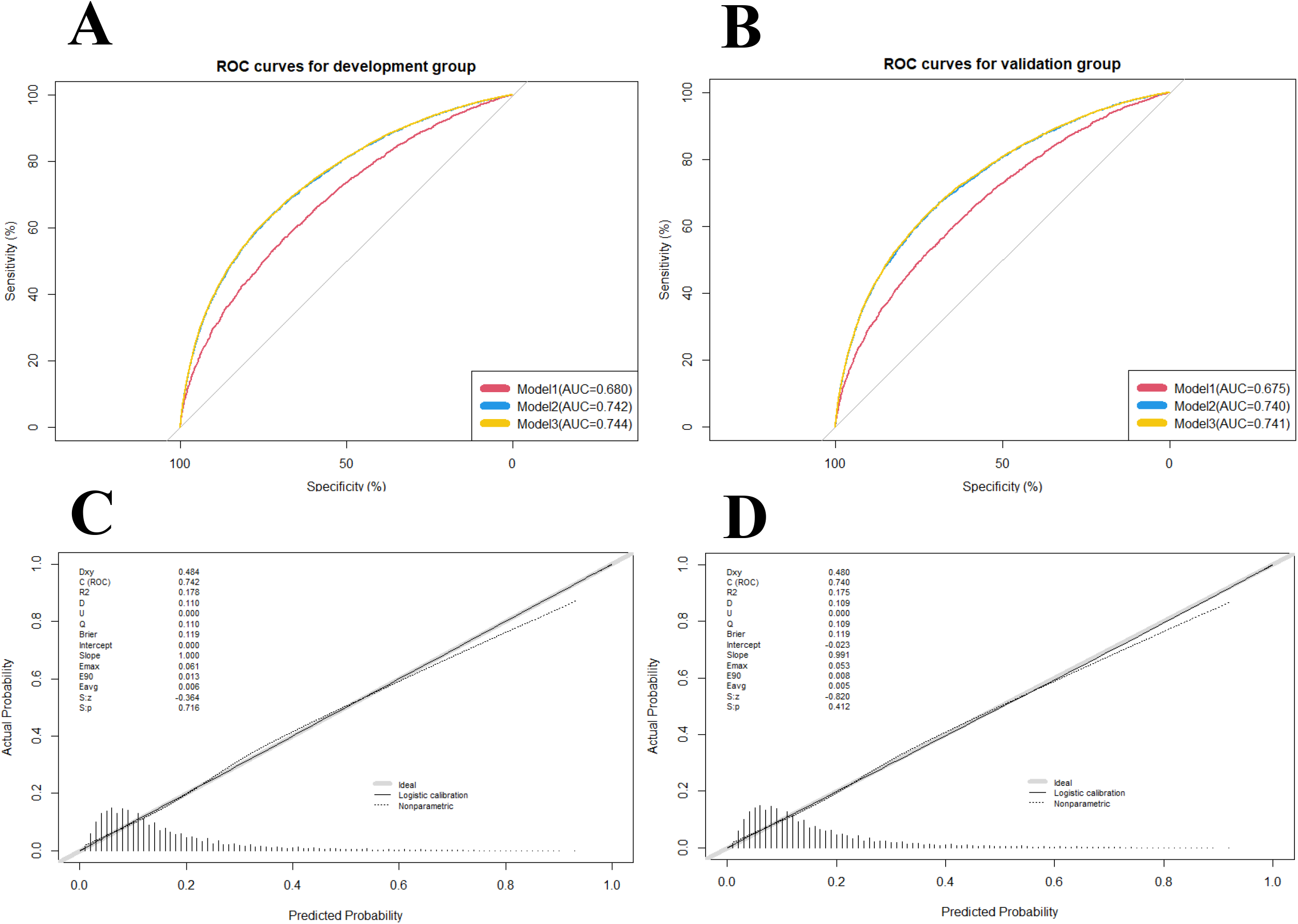
Receiver operating characteristic (ROC) curves for the prediction of high blood pressure in the training group and validation group. (A) ROC curves of the factors and nomogram in the development group; (B) ROC curves of the factors and nomogram in the training group; (C) Calibration plot of nomogram prediction in the development group; (D) Calibration plot of nomogram prediction in the validation group. ROC curves from the prediction model and other predictive strategies(Model1:Age, gender, gestational hypertension, weight status, family history of hypertension, family history of obesity, average outdoor physical activity time; Model2: Age, gender, gestational hypertension, weight status, family history of hypertension, family history of obesity, average outdoor physical activity time, birthweight, feeding mode; Model3: Age, gender, gestational hypertension, weight status, family history of hypertension, family history of obesity, average outdoor physical activity time, birthweight, feeding mode; parental smoking status, parental education level, household monthly income, average screen-based time, fried food intake.) for comparison. Calibration curve represents the calibration of the nomogram, which shows the consistency between the predicted probability of conversion and actual conversion probability of HBP patients. The x-axis is the predicted probability by nomogram and the y-axis is the actual conversion rate of HBP patients. The grey line represents a perfect prediction by an ideal model, and the black-dotted line shows the performance of the nomogram, of which a closer fit to the grey line means a better prediction.

**Figure.3.**
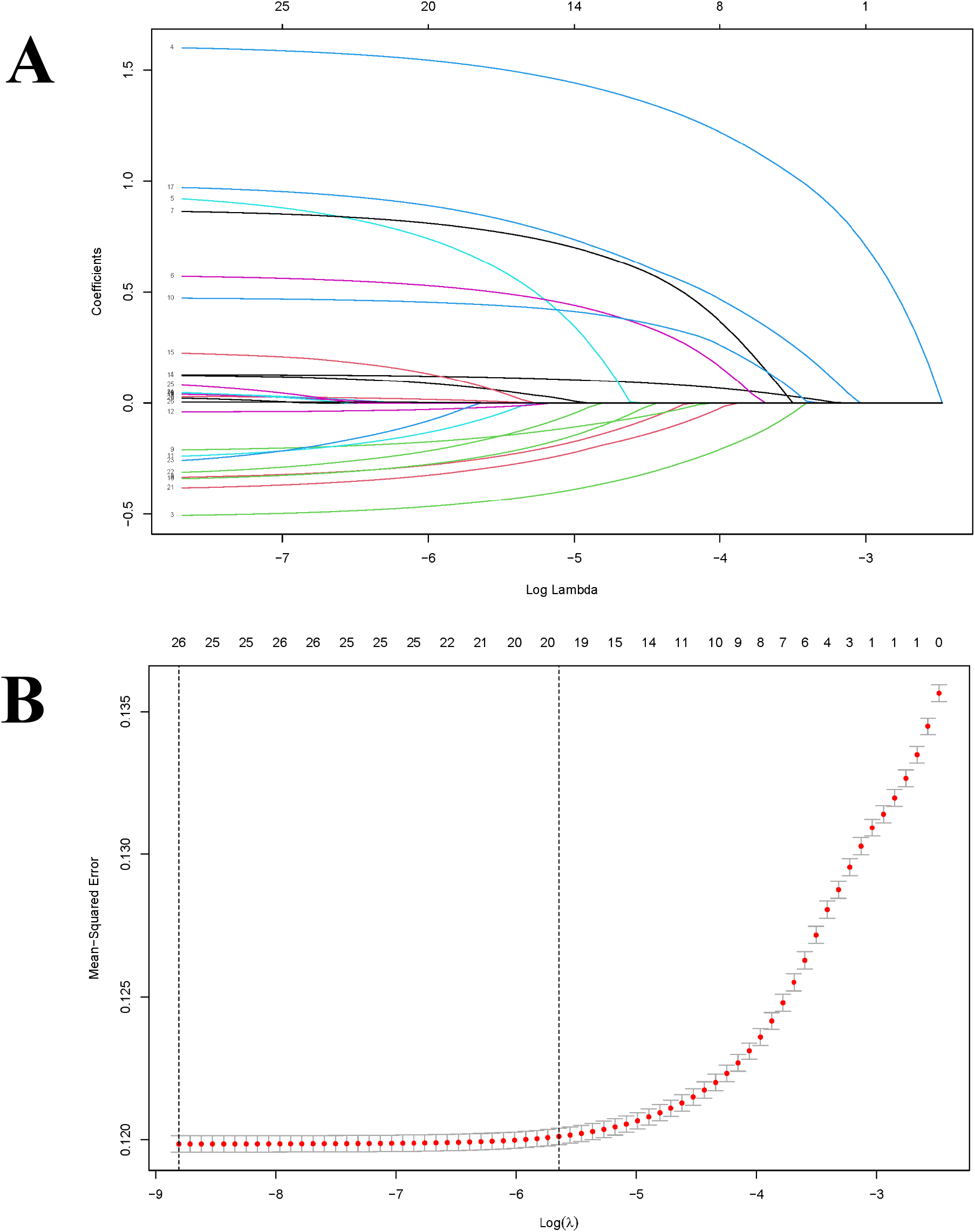
Predictor selection using the LASSO binary logistic regression model. A LASSO coefficient of the total 14 predictors. (A) LASSO coefficient profiles of all predictors, a coefficient profile plot was provided against the log (Lambda) sequence. (B) Predictors selection by LASSO via minimum criteria, predictor selection in the LASSO model used tenfold cross-validation via minimum criteria. Red-dotted vertical lines were drawn at the optimal values by using the minimum criteria (minimize the mean-squared error), the value 9 represents those 14 predictors were reduced to 9 nonzero features by LASSO.

### Website of nomogram

An available web-based nomogram calculator of HBP was built to present the diagnostic probability for helping guardians and physicians to identify the HBP among children and adolescents in a user-friendly way.

## Discussion

As the high potential hazard predictors reported previously, age, gender, birthweight, feeding mode, gestational hypertension, weight status, family history of obesity, family history of hypertension and frequency of PA were also be found and presented as a simple and reliable nomogram plot which designed for identifying the children and adolescents who were at high risk of developing HBP.

Accumulating evidence supports the view that the roots contribution of essential hypertension extends back to childhood and adolescence(Cosenzi, S Ac Erdote, Bocin, Molino, & Bellini, 1996; Kawabe et al., 2000). Targeted identifying the risk factors of developing HBP in children and adolescents may be practicable to avoid unnecessary burden, overdiagnosis or costs. Our study provides a more interpretability, more precise classification standard for HBP in youths, instead of using complicated model to predict HBP.

The strongest risk factor for primary HBP in children and adolescents is elevated body mass index(Giussani et al., 2013), which was also demonstrated in our study. A higher prevalence in children with HBP who were classified as overweight, was observed in the comprehensive meta-analysis(Song, Zhang, Yu, Zha, & Rudan, 2019). Similar prevalence values were also noted in previous studies conducted in American and European suggest that the incidence of HBP among children and adolescents with obesity had significantly increased compared to the non-obese children(Chiolero et al., 2007; Kaelber et al., 2016). There are several known pathophysiological pathways by which obesity may lead to an elevated BP and HBP. A central tent for HBP is that dysfunctional adipocyte and neurohormonal activation of the sympathetic nervous system(SNS). Increased SNS activity can leads to elevated BP and HBP by increasing renin–angiotensin–aldosterone system (RAAS) activity in addition to the direct vasoconstricting effects of SNS. RAAS activity increases BP directly (angiotensin II-mediated vasoconstriction and further SNS activation) and indirectly (angiotensin II- and aldosterone-mediated salt and water tubular reabsorption and ADH-mediated water retention)(Brady, 2017; Lagos et al., 2016). Additionally, because of that kidney would become fat encapsulated with the obsess state, there is increased renal interstitial fluid pressure and slower tubular flow rates which ultimately leads to increased sodium reabsorption and increased intravascular volume(Ouchi, Parker, Lugus, & Walsh), which may cause the hypertension occurs. Evidence is accumulating that factors during early-life stage have long-term effects on later BP in childhood and young adulthood(Barker & Djp, 2010; Cohen & Meryl, 2004). As the indicator operating early in life, history of low or high birthweight(birthweight<2500g or >4000g) seemed lead to be associated with a higher risk of HBP(Bergvall, Iliadou, Tuvemo, & Cnattingius, 2005; Gansäuer, Rosales, & Justicia, 2005). Nevertheless, some studies did not support this association(Daly, Scragg, Schaaf, & Metcalf, 2005). With regard another modifiable environmental factors, previous studies that corroborated the protective role of breastfeeding on BP later in life(Kelishadi et al., 2006; Martin & R, 2005), on the contrary, our model indicate that compared with breastfeeding mode, partial breastfeeding may play a protective role while bottle feeding may considered to be risk role to individuals, another meta-analysis indicate that any effect of breastfeeding on BP is modest with limited public health importance may partly attribute to the publication bias(Owen et al., 2003). Notwithstanding the early-life factors were possible associations with increased risk for HBP among youths is inconclusive, their long-term efficacy raises concerns and further investigation are desirable.

Our findings, which were consistent with majority of studies, revealed that other risk factors were increased the risk of HBP, which including male sex, and family history of hypertension, as well as insufficient of exercise(Hamoen et al., 2020; Li, Sun, Liu, Li, & Zhou, 2019; Moyer & Virginia, 2013). Our model shows that lower levels of PA is correlated with higher risk of HBP, and other studies have commonly shown a beneficial role of PA for youths with HBP(Strong et al., 2005). Moreover, our finding also suggested that an excess of Sedentary behavior time(SBT) is associated with an increased risk of hypertension compared to the recommended levels of SBT. Overall, efforts to encourage youths increase their PA, and reduce their time of SBT are warranted. For fired food intake, based on the initial regression shrinkage and selection of Lasso, there were no statistically significant differences in youths for hypertension. This discrepancy could be linked to the fact that salt intake in this study did not include due to the lack of data, even though we use fired food instead of salt intake roughly.

Previous studies using a similar methodology supported the hypothesis that age, gender, birthweight, family history of HBP and other predictors may account for the high prevalence of HBP during childhood and adolescence. Nevertheless, other models suggested that child ethnicity, environmental and genetic risk factors play critical roles in onset and progression of HBP(Ataei et al., 2004; Hamoen et al., 2020). It should be noted that the regional differences, eating habits and differences morphometric characteristic can explain the dissimilar findings between studies(El-Shafie, El-Gendy, Allhony, Fotoh, & Galab, 2018). Of the examined risk factors, both age and gender are important contributions to the progression of HBP, and may be more significant if other modifiable risk factors were involved in their life, which is particularly true for essential hypertension(Antal et al., 2004). Since dichotomization of continuous variables may cause loss of valuable information(Royston, Altman, & Sauerbrei, 2005; Steyerberg & Vergouwe, 2014), we have categorized continuous predictors such as PA and SBT, especially weight status(BMI), because of normal changes in BMI that occur as children age(Giussani et al., 2013).

To date, our study has established a quantitative nomogram for the first time in the form of a concise, more practical, policy-making orientation, HBP nomogram for children that not only useful for clinicians, public health practitioners and policymaker in the developed countries to address this global health need among various indications but also, and perhaps more importance, for those in the low-income countries. In the point of macro-view, identifying the HBP predictors for youths would not only relieving the burden of monitor and administer on youth health, but also for economizing the facilities and resources. Furthermore, such identifications can help introducing earlier interventions for those at risk, which include, but are not limited to, help children disengage themselves from the concern of diseases early as possible, and prevent the transfer to other potential hazards such as adverse cardiovascular outcomes. In the methodological point of view, numerous selective variables were incorporate into a comprehensive instrument for the prediction of the probability of new high blood disorder among youth. Summary variables selected were based on the individuals’ demographic characteristics, family history, physical examination and lifestyle-associated factors. To improved approach avoided the concerns of model overfitting, the predictive value of the nomogram is easy to handle, which was evaluated based on highly discrimination, calibration, and clinical utility in separate internal training and validation datasets. One recent publication has developed and validated a risk prediction model for screening the hypertension among Chinese adults(Deng et al., 2021). Targeting on the large-scale study on youth population, our study suggest that 9 predictors are significantly associated with HBP among youths. All predictors are readily available in routine physical examinations and easily measured by individuals themselves, which means the established model in our study represent a possibly useful instrument for rapid assessment of hypertension among children and adolescents compared with the former model. Furthermore, our study has more advantages than another prediction model based on the youth population with a higher discrimination, larger sample size and more selective candidates for estimating simultaneously(Hamoen et al., 2020).

However, several limitations exist in the present study. We developed a nomogram model for estimating the risk of HBP while validated it with an internal assessment dataset. The incidence of HBP (16.9%) in our study was higher than the reported incidence in the general youths(4.0%)(Song et al., 2019), which may have biased our results. More characteristics of patients and variables should be established and extracted to improve the capacity of predicting prognosis in youths with HBP, both genetic and environmental determinants are likely to be responsible for the HBP occurs(Li et al., 2019).

## Conclusion

In this study, a feasible nomogram was derived from the ongoing study to visually predict the probability of HBP in children and adolescents for clinicians, policy-maker even the family members on enhancement of the screening and subsequent diagnosis.

## Methods

### Study population and Data collection

The survey is not publicly available and participants were protected under a certificate of confidentiality issued by the Government of Guangzhou due to the sensitivity nature of data collected from all students group in Guangzhou city.

Our study was performed in accordance with the Strengthening the Reporting of Observational Studies in Epidemiology(STROBE) reporting guideline(Von, Altman, & Egger, 2007). Relevant information was collected from all participants’ parents or their legal guardians gave written informed consent which performed according to the guidelines of the Declaration of Helsinki(Association, 2009).

This survey was approved by the Ethics and Human Subject Committee of Sun Yat-Sen University and any questions were reach a consensus before the consent forms were signed. Data were obtained from the GSSCH, a sequential cross-sectional investigation carried out annually in comprised approximately 1,600 primary and middle schools in Guangzhou city, which done and supported by Sun-Yat Sen university, Guangzhou school-health promotion center and Guangzhou Education Bureau. The GSSCH has been conducted for 4 consecutive years, and battery of relevant measures were done during September and October, the first two months of a new school year.

### Collecting data and Definition of covariates

The dataset was independently divided by the cross-sectional study conducted in 2020 for identifying the first-ever incidence of HBP among children and adolescents. All participants were students aged from 5 to 25 years old, and they were examined by trained investigators or practitioners for questionnaires on health-related behaviors, regular physical examinations, as well as objectively anthropometric data measurements. 1,102,462 children were removed imputation of missing data, incomplete data, or the extreme values respecting to the demographic variables or relevant BP variables based on the 4 Standard deviation(SD) of sex-and age-specific. 148,850 participants were aged above 18. 342,736 subjects were included in our analysis after the above-mentioned eliminations. The subject group was split into two parts, training and validation sets, by taking a random sample of 70%(N=239,546) of the training and the remaining 30%(N=103,190) for validation.

Variable information comprised the demographic characteristics (e.g., age, sex, height, weight, family income), lifestyle-associated factors [e.g., Physical activity(PA), dietary pattern, sleep pattern] and health-related information(including medical history, past, current diseases and family history) for hypertension were assessed in 2020 as the training cohort and validation database, through matching individuals’ identifications. Body mass index(BMI) was calculated as the individuals body weight in kilograms divided by the square of height in meters, and further divided into four categories (underweight, normal weight, overweight, obese) adjusted for age and gender specific. Parental education level was defined as the highest level of education reported for either the parents, which was divided into, primary school or below, secondary school, senior high school or junior school, junior college or university, graduate or above. Parental smoking status separately for the father and mother was combined with three categories, namely, never smokers (both parents were never smokers), former, or current smokers (either of the parents were former or current smokers). The family history of obesity and hypertension was defined by reported history of obesity and hypertension in immediate family members, while the gestational hypertension was defined as new-onset hypertension after 20 weeks’ gestation.

### Measurement of High Blood pressure

The child was requested for seating comfortably at least 10 minutes or longer before BP measurement which was conducted in a quiet platform thenceforth. A standard BP cuff was positioned nearby the right arm elbow, 2 centimeters below the antecubital fossa, and BP for each child was recorded 3 consecutive times in seated position on their right arm by a calibrated electronic sphygmomanometer (OMRON, HEM-4001C, Kyoto, Japan) in single occasion. The diagnosis of HBP was in accordance with the updating BP diagnostic criteria for Chinese children and adolescents that average Systolic blood pressure(SBP) or Diastolic blood pressure(DBP) were greater than or equal to the 95^th^ percentile for the corresponding of child’s age, sex and height(Fan, Yan, Jie, & Epidemiology, 2017).

### Statistical analyses

The continuous variables of participants were presented as mean with standard deviation and interquartile range (IQR), whereas categorical data were revealed as absolute frequencies with percentages. For differences between groups of continuous variables and categorical, the two-sample *t* test or Mann-Whitney test and Chi-Square test were applied, as appropriate. We first used LASSO-penalized logistic regression to identify significant predictors by presenting the significance with α=0.05 with over 1000 bootstrap iterations(Tibshirani, 2011). The Odds Ratio(OR) with the following 95% Confident Interval(CI) were computed as well. Multivariable logistic regression was generated by putting consecutive candidate variable factors which discard the predictors according to the *P*-values with the stepwise manner(E. W. S. A et al., 2001). Along with the nomogram-predicted probability of HBP was constructed. The Bayesian information criterion and Akaike information criterion(AIC) were used for evaluating the maximum likelihood model respecting to multivariable logistic regression. Moreover, we constructed the Area under the receiver-operating characteristic curve(AUC, or C-index) to assessed the model discrimination while Hosmer-Lemeshow approach and calibration curve was built for detecting model calibration. Here, all analyses were conducted using the R language (X64 Version 4.1.0, R Foundation for Statistical Computing, Vienna, Austria, https://www.r-project.org/) with several packages including “Foreign”, “Hmisc”, “Glmnet”, “Caret”, “Rms” and “pROC”. The bilateral *P*<0.05 was recognized as statistically significant.

## Data Availability

The survey is not publicly available and participants were protected under a certificate of confidentiality issued by the Government of Guangzhou due to the sensitivity nature of data collected from all students group in Guangzhou city. Requests to assess the dataset from qualified researchers trained in human participant confidentiality protocols may be sent to the School of public Health, Medical College of Sun Yat-Sen University at.

## Conflict of interest

All other authors declare that they have no conflicts of interest.

---

**Supplementary Figure.1.**
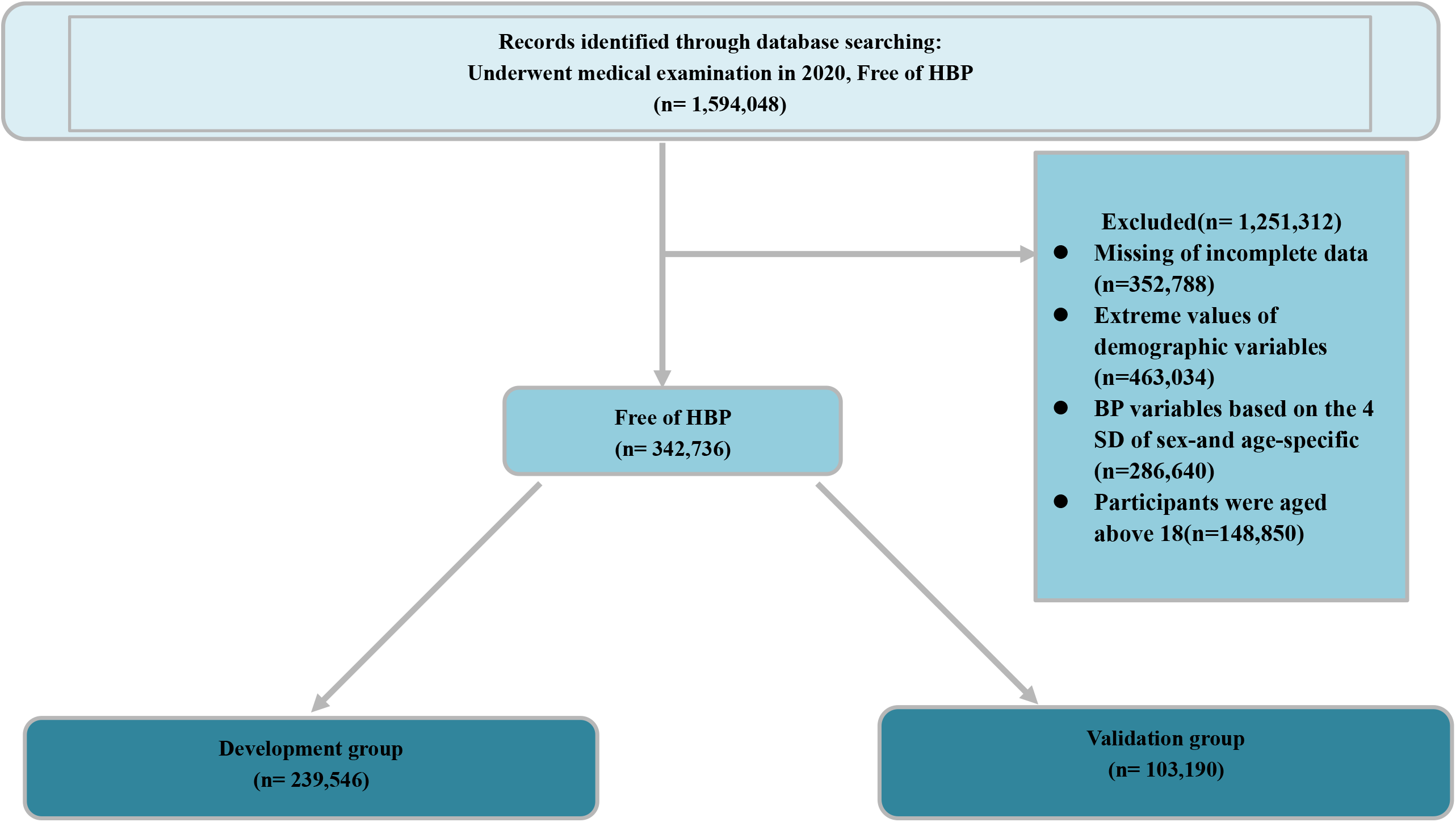
Flow chart illustrating the inclusion and exclusion of participants into the study. BP, Blood pressure; HBP, High blood pressure; SD, Standard deviation.

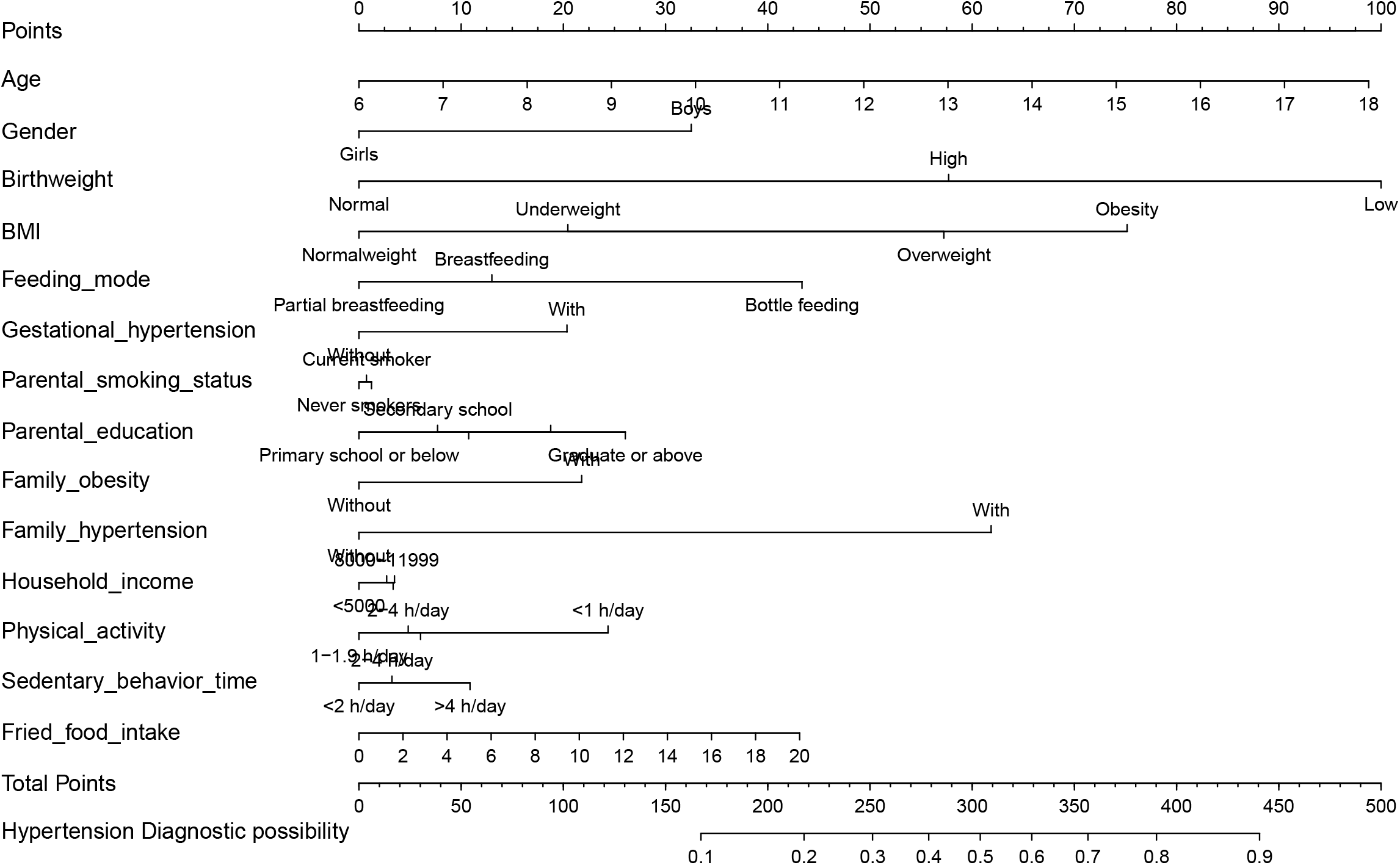

**Supplementary Figure.3.**
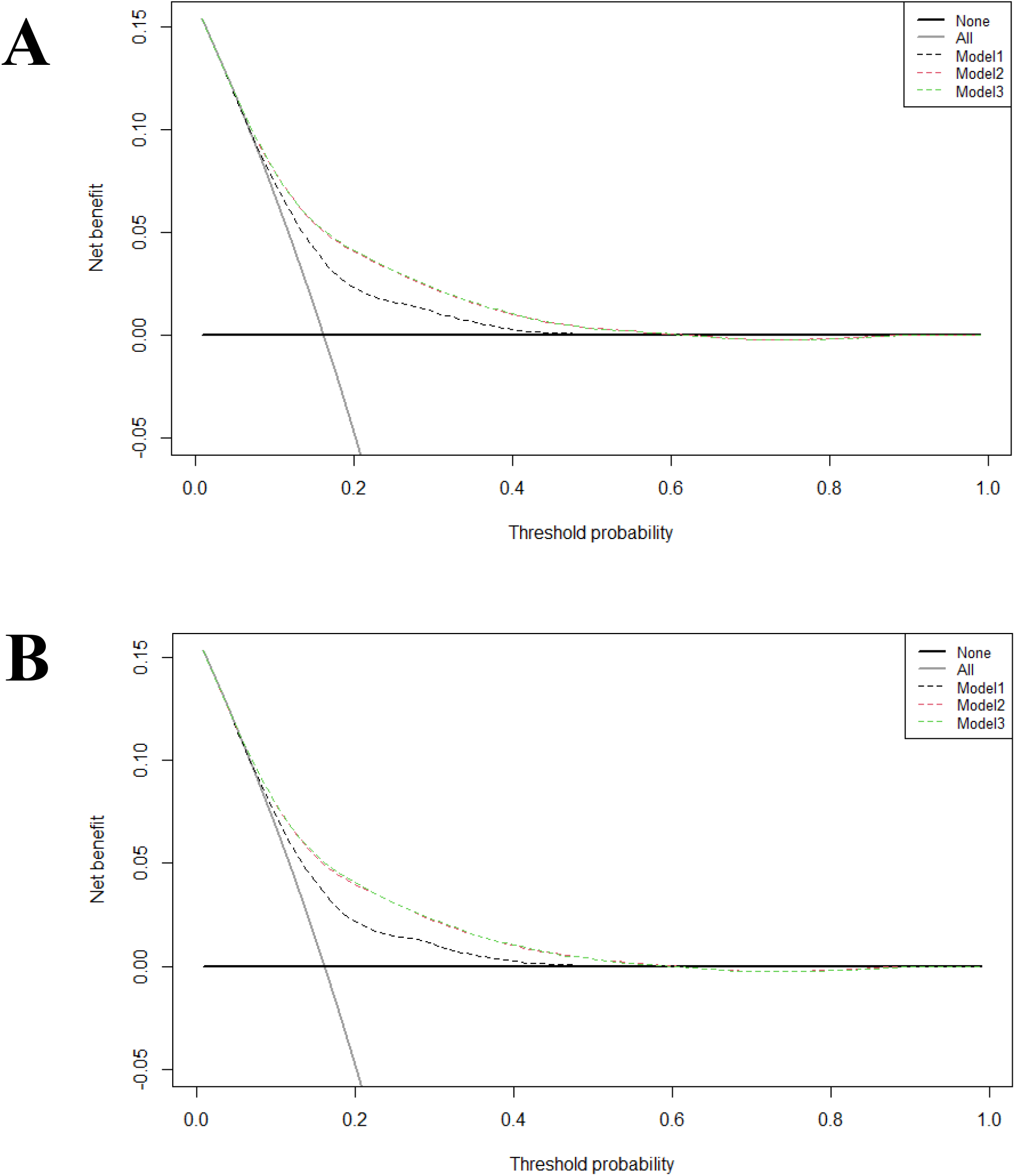
Decision curve analysis of the nomogram prediction in the training group and validation group. (A) Decision curve analysis of the nomogram prediction in the training group; (B) Decision curve analysis of the nomogram prediction in the validation group; Model1:Age, gender, gestational hypertension, weight status, family history of hypertension, family history of obesity, average outdoor physical activity time; Model2: Age, gender, gestational hypertension, weight status, family history of hypertension, family history of obesity, average outdoor physical activity time, birthweight, feeding mode; Model3: Age, gender, gestational hypertension, weight status, family history of hypertension, family history of obesity, average outdoor physical activity time, birthweight, feeding mode; parental smoking status, parental education level, household monthly income, average screen-based time, fried food intake.

## Notes

### Competing Interest Statement

The authors have declared no competing interest.

### Clinical Trial

Not required.

### Author Declarations

This survey was approved by the Ethics and Human Subject Committee of Sun Yat-Sen University and any questions were reach a consensus before the consent forms were signed.

